# Epidemiological Risk Factors Associated with Death and Severe Disease in Patients Suffering From COVID-19: A Comprehensive Systematic Review and Meta-analysis

**DOI:** 10.1101/2020.06.19.20135483

**Authors:** Kunchok Dorjee, Hyunju Kim

## Abstract

**Introduction:** Progression of COVID-19 to severe disease and death is insufficiently understood.

**Objective:** Summarize the prevalence adverse outcomes, risk factors, and association of risk factors with adverse outcomes in COVID-19 patients.

**Methods:** We searched Medline, Embase and Web of Science for case-series and observational studies of hospitalized COVID-19 patients through May 22, 2020. Data were analyzed by fixed-effects meta-analysis, using Shore’s adjusted confidence intervals to address heterogeneity.

**Results:** Forty-four studies comprising 20594 hospitalized patients met inclusion criteria; 12591 from the US-Europe and 7885 from China. Pooled prevalence of death [%(95% CI)] was 18% (15-22%). Of those that died, 76% were aged≥ 60 years, 68% were males, and 63%, 38%, and 29% had hypertension, diabetes and heart disease, respectively. The case fatality risk [%(95% CI)] were 62% (48-78) for heart disease, 51% (36-71) for COPD, and 42% (34-50) for age≥ 60 years and 49% (33-71) for chronic kidney disease (CKD). Summary relative risk (sRR) of death were higher for age≥ 60 years [sRR=3.8; 95% CI: 2.9-4.8; n=12 studies], males [1.3; 1.2-1.5; 17], smoking history [1.9; 1.1-3.3; n=6], COPD [2.0; 1.6-2.4; n=9], hypertension [1.8; 1.7-2.0; n=14], diabetes [1.5; 1.4-1.7; n=16], heart disease [2.0; 1.7-2.4; 16] and CKD [2.0; 1.3-3.1; 8]. The overall prevalence of hypertension (55%), diabetes (31%) and heart disease (16%) among COVODI-19 patients in the US were substantially higher than the general US population.

**Conclusions:** Public health screening for COVID-19 can be prioritized based on risk-groups. A higher prevalence of cardiovascular risk factors in COVID-19 patients can suggest increased risk of SARS-CoV-2 acquisition in the population.

## Introduction

Coronavirus disease-19 (COVID-19) caused by severe acute respiratory syndrome-coronavirus-2 (SARS-CoV-2) that first emerged in Wuhan, China in late December 2019 has spread with such rapidity and efficiency that in less than six months, it has caused more than 8.5 million cases and 450,000 deaths globally.^1^ Driven by an urgency to solve the crisis, studies are being published at an unprecedented pace. However, across the publications, prevalence of death, severe disease and their association with epidemiological risk factors have greatly varied^2,3^, with studies showing conflicting results for association of key risk factors such as sex,^4-8^ smoking,^9-12^ hypertension^5,6,8,13,14^ and diabetes^5,6,8,13,14^ with COVID-19 disease severity and death. Whether or how cardiovascular risk factors, especially prior hypertension, diabetes and heart disease are associated with acquisition of SARS-CoV-2 and progression to severe disease or death is being debated and not understood well.^15-17^ Meta-analyses conducted so far on prevalence of epidemiological risk factors and association with disease progression were mostly based on studies from China^16,18,19^ and many of the analyses on prevalence estimates included studies focused on critically ill patients,^9,18,20,21^ which can overestimate the prevalence and affect generalizability of results. To our knowledge, none of the analyses were restricted to hospitalized COVID-19 patients that indeed provides an efficient sampling frame to investigate disease progression in relation to risk factors.

Therefore, we undertook a comprehensive systematic review and meta-analysis to investigate the association between key epidemiological factors–age, gender, smoking, hypertension, diabetes, heart disease, chronic obstructive pulmonary disease (COPD), and chronic kidney disease (CKD)–and progression to death in patients hospitalized due to COVID-19. We additionally compared the 1) the prevalence of risk factors and death in the US-Europe with that of China; 2) the prevalence of co-morbidities at baseline with the general population prevalence, and 3) prevalence of cardiovascular disease, chronic kidney disease and COPD at baseline with corresponding organ injuries (acute cardiac injury, acute lung injury, and acute kidney injury) during hospital admission.

## Methods

### Literature search, study selection and data abstraction

We searched Medline, Embase, Web of Science and the WHO COVID-19 database to identify studies published through May 22, 2020 that investigated the risk of severe disease or death in hospitalized patients with confirmed COVID-19 disease. We used search terms, ‘coronavirus disease 19’, ‘COVID-19’, ‘severe acute respiratory syndrome coronavirus 2’ and ‘SARS-CoV-2’. We started the search on March 18, 2020 with biweekly search thereafter and final search on May 22, 2020. We included case series and observational studies that described the prevalence of death or severe disease in adult population stratified by risk factors: age, sex, hypertension, diabetes, heart disease, COPD, CKD and CLD. We excluded studies that exclusively focused on pregnant women, children, and elderly patients. We excluded studies that exclusively studied critically ill patients from calculation of prevalence of death but included them for calculating the association of risk factors with death.

### Risk factors and outcomes

Primary outcome was prevalence of death and association of risk factors with death. We extracted data on death as recorded in the publications. We measured prevalence of severe disease and association with risk factors as secondary outcome. We defined outcome as severe disease for any of 1) the study classified COVID-19 disease as severe or critical, 2) intensive care unit (ICU) admission, 3) acute respiratory distress syndrome, or 4) mechanical ventilation. Severe disease was defined by studies as respiratory rate≥ 30 per minute, oxygen saturation≤ 93%, and PaO_2_/FiO_2_ <300 and/or lung infiltrates>50% within 24-48 hours.^3^ Critical illness was defined as respiratory failure, shock and/or multiple organ dysfunction or failure.^3^ Heart disease as a pre-existing condition was broadly defined by most studies as ‘cardiovascular disease’ (CVD). Additional outcomes were acute cardiac and and kidney injury in the hospitalized patients that were defined as such by the studies.

### Statistical analysis

We calculated and reported summary estimates from fixed-effects models ^22^. We assessed heterogeneity across studies using Cochran’s Q-test (*η*^2^ p value <0.10) ^23^ and I^2^ statistics (I^2^ >30%) ^24^. In the presence of heterogeneity, we adjusted the 95% confidence intervals for between-study heterogeneity using the method described by Shore et al ^25^. We have presented the results from random effects meta-analysis as well. The meta-analysis was performed in Microsoft^®^ Excel 2020 (Microsoft Corporation, Redmond, WA). We analyzed publication bias using funnel plots, and Begg’s and Egger’s tests. Quality of each study was assessed using the Newcastle-Ottawa assessment scales using the PRISMA guidelines. We calculated the following as a part of our analyses: 1) prevalence of severe disease or death, 2) prevalence of risk factors, and 3) relative risk for the association of age, sex, and comorbidities with outcome. When not reported or when unadjusted odds ratio was presented, we calculated the relative risk (95% confidence interval) using the frequencies provided. Adjusted estimates were used where available. Case fatality risk (and case severity risk) for a specific risk factor was calculated as number of deaths (or severe disease) in patients with a risk factor out of all patients possessing the risk factor.

## Results

### Study characteristics

Initial search yielded 13919 citations. Articles were then filtered (**Figure 1**). We identified 275 articles for full text review, of which 44 studies met inclusion criteria (**Table 1**).^4,5,7,8,13,14,26-61^ The studies were conducted in: China (n=31), USA (n=8), Italy (n=2), UK (n=1), Iran (n=1) and Singapore (n=1). Two studies were prospective, one cross sectional, and remaining retrospective in nature.

**Table 1.**
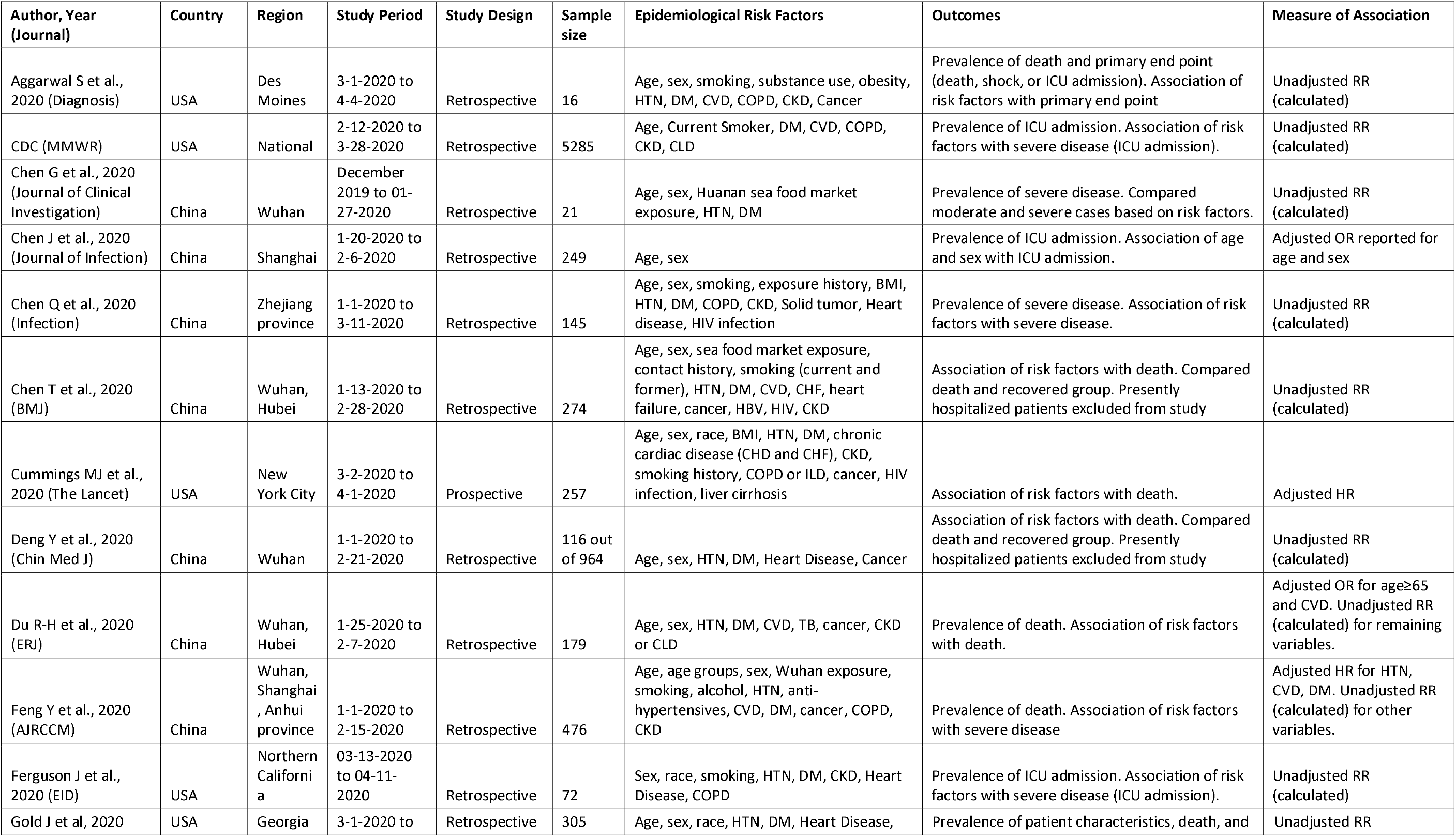
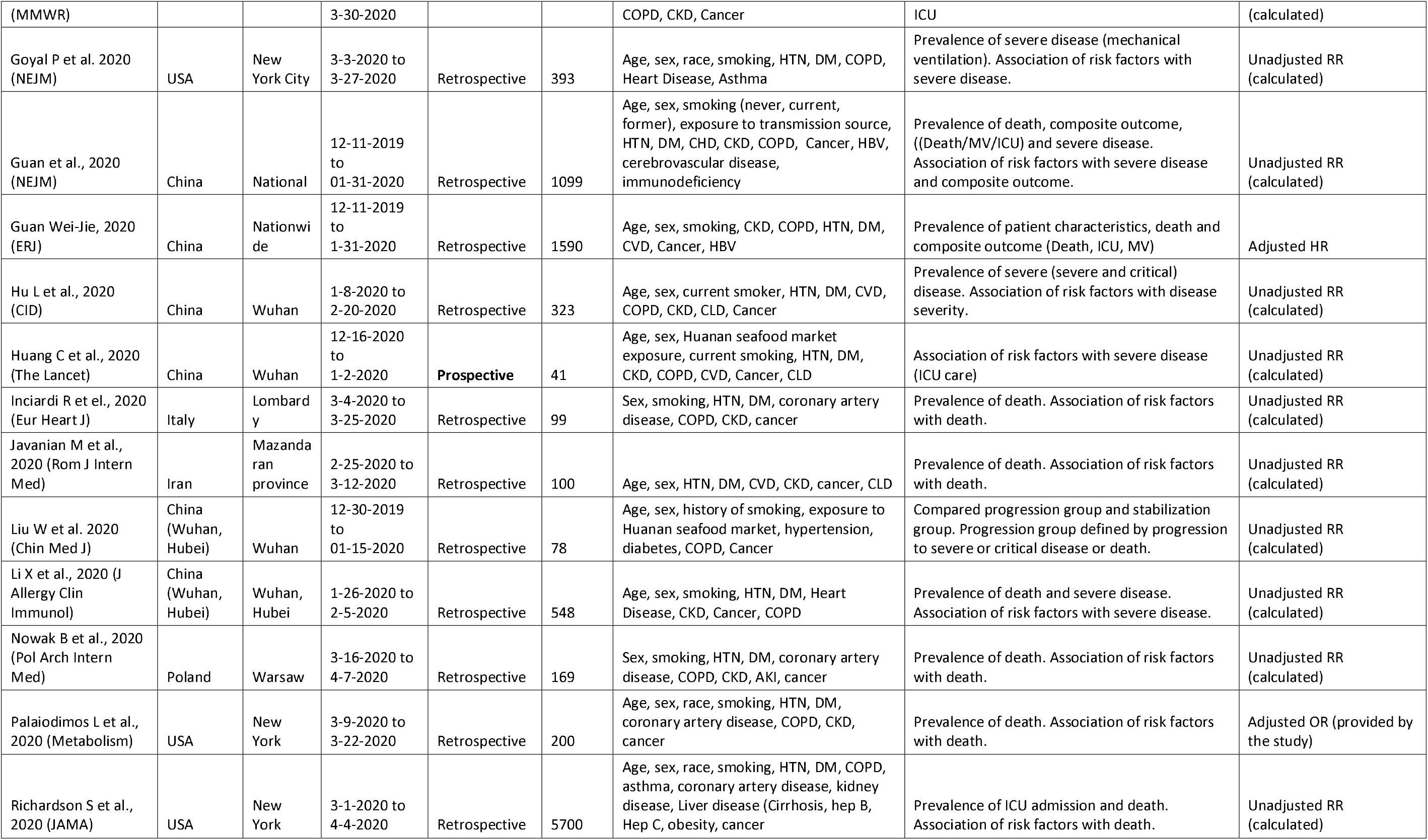
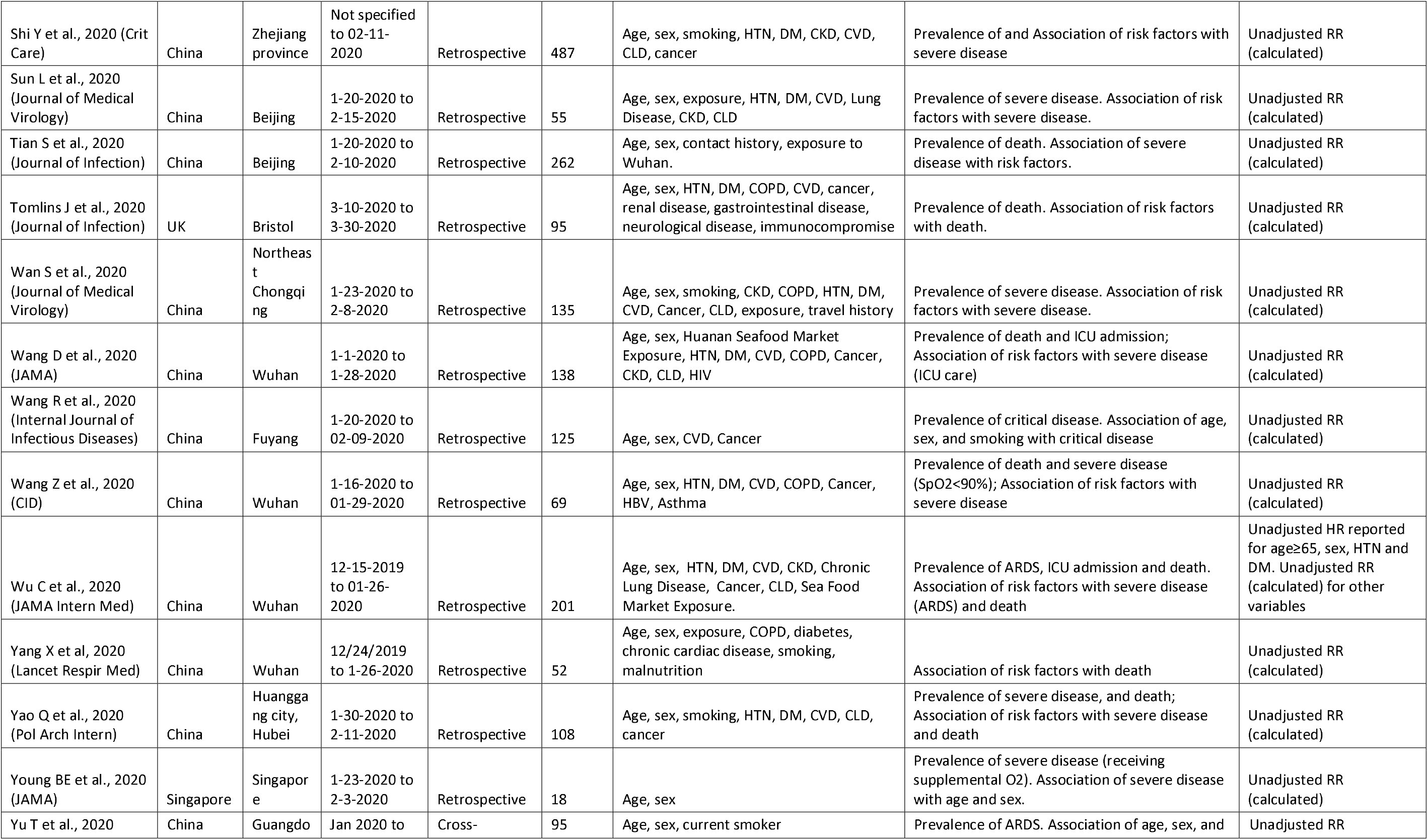
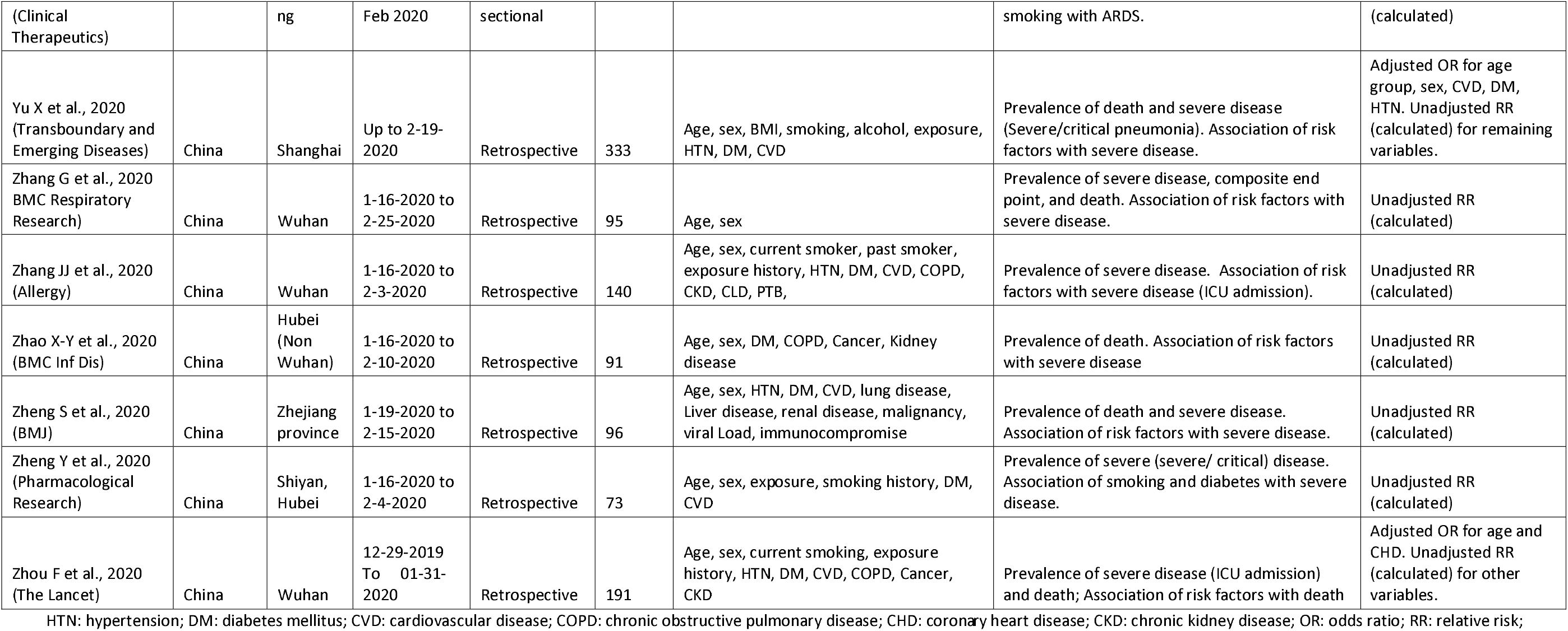
Characteristics of studies in COVID-19 hospitalized patients to investigate association of risk factors with death or severe disease

**Figure 1.**
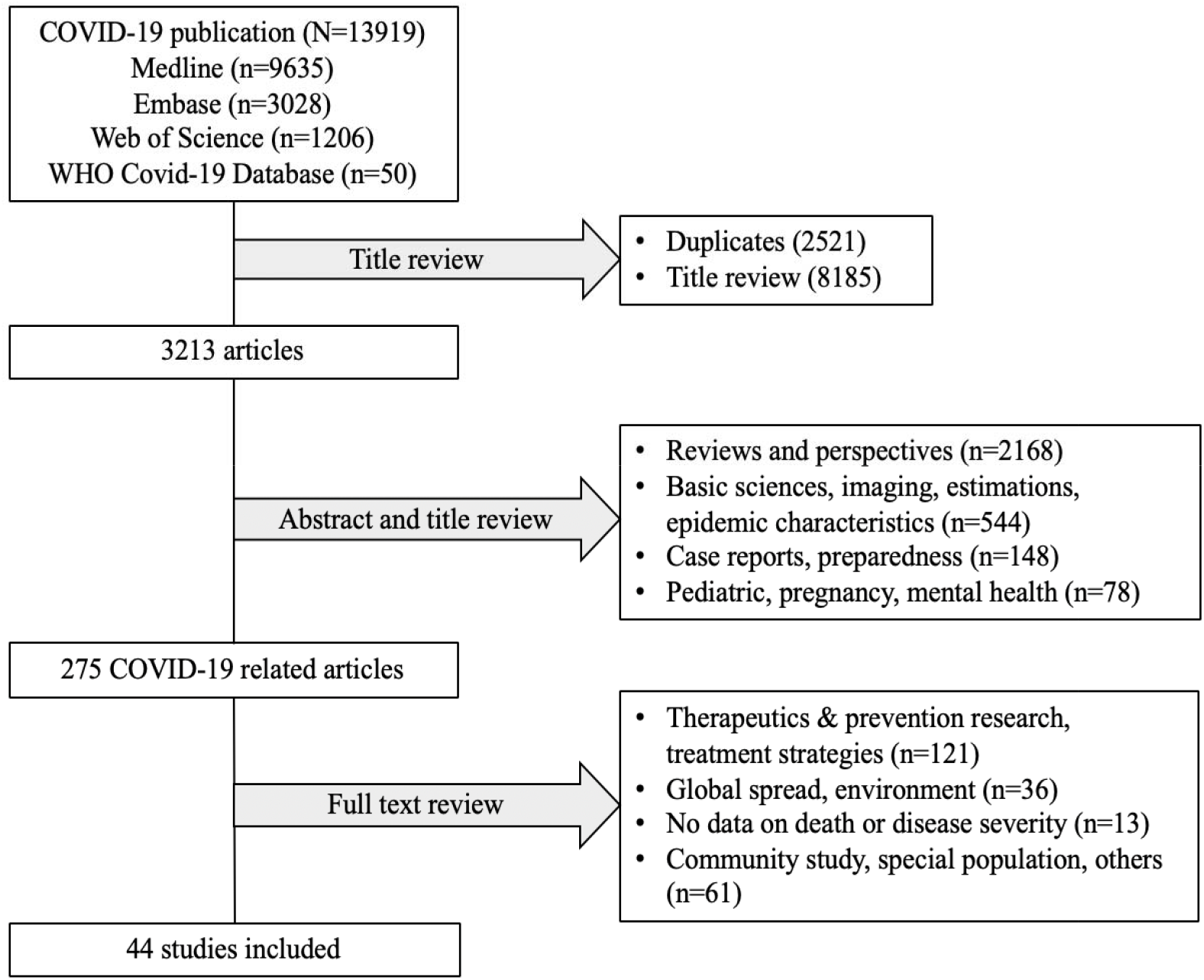
Identification of studies.

### Population and demographics

There were 20594 total COVID-19 hospitalized patients that included 12591 patients from the US and Europe (>90% from the US), and 7885 patients from China. Median age was 57 years [IQR: 54-60 years; I^2^=62%; n=36 studies]; 65 years [IQR: 61-70 years; I^2^=0%; n=9] for the US and Europe; and 54 years [IQR: 51-58 years; I^2^=0%; n=26] for China. Forty-three percent [95% CI: 38-48%; I^2^=96%; n=22] were aged≥ 60 (**Table 2**); 46% [95% CI: 43-50%; I^2^=88%; n=4] for US and Europe; and 31% [95% CI: 25-38%; I^2^=95%; n=18] for China. Fifty-eight percent [(95% CI: 56-60%); I^2^=78%; n=43] were males.

**Table 2.** Pooled prevalence of death and severe disease stratified by epidemiological risk factors in COVID-19 patients hospitalized between December 2019-May 2020

| Risk group | Overall prevalence of risk across studies |  | Pooled Prevalence of Death (Case Fatality Risk) and Risk Factor |  |  | Summary Relative Risk of Death |  |  |  |
| --- | --- | --- | --- | --- | --- | --- | --- | --- | --- |
| | No. of studies | Pooled prevalence of risk factor, % (95% CI) | No. of studies | *Case fatality risk (Pooled prevalence of death in risk group), % (95% CI) | #Prevalence of risk group in dead, % (95% CI) | No. of studies | Fixed Effects<br>Summary relative risk; 95% CI (Shore adjusted) | Random Effects<sup>#</sup><br>sRR; (95% CI) | Heterogeneity<br>$I^2$; $\chi^2$; p value |
| Age ≥ 60 years | 22 | 43 (38-48) | 8 | 42 (34-50) | 76 (71-82) | 12 | 3.77 (2.94-4.82) | 1.66 (1.21-2.27) | 73%; 41; p<0.01 |
| Male | 43 | 58 (56-60) | 16 | 30 (23-38) | 68 (63-74) | 17 | 1.34 (1.20-1.50) | 1.39 (1.22-1.58) | 19%; 16; p=0.20 |
| Smoking history | 22 | 13 (11-16) | 5 | 22 (12-38) | 27 (18-41) | 6 | 1.87 (1.05-3.33) | 1.89 (1.03-3.44) | 80%; 25; p<0.01 |
| Current smoker | 13 | 8 (6-12) | 3 | 21 (8-56) | 21 (13-37) | 4 | 2.20 (1.16-4.16) | 2.51 (1.30-4.86) | 78%; 14; p<0.01 |
| COPD | 27 | 8 (6-10) | 8 | 51 (36-71) | 12 (7-21) | 9 | 1.97 (1.59-2.43) | homogeneous | 0%; 3.55; p=0.83 |
| Hypertension | 34 | 47 (41-54) | 13 | 24 (16-36) | 63 (57-70) | 14 | 1.84 (1.66-2.03) | homogenous | 34%; 20; p=0.10 |
| Diabetes | 36 | 26 (23-30) | 13 | 22 (16-32) | 38 (32-44) | 16 | 1.51 (1.37-1.68) | homogeneous | 0%; 10; p=0.68 |
| Cardiovascular disease | 35 | 14 (12-17) | 12 | 62 (48-78) | 29 (21-41) | 16 | 1.99 (1.72-2.38) | homogenous | 33% (22; p=0.10 |
| Chronic kidney disease | 26 | 7 (5-10) | 7 | 49 (33-71) | 23 (13-39) | 8 | 2.02 (1.30-3.13) | 2.14 (1.31-3.51) | 75%; 28; p<0.01 |

\* Case fatality risk of represent total number of people that died in the specific risk group divided by total population in the risk group.
<sup>#</sup> Prevalence of risk group in dead represent total number of people having the risk group divided by total population that died.

### Prevalence of Death and Severe Disease

We calculated a pooled prevalence of death of 18% [95% CI: 15-22%; I^2^=92%; n=26], ranging from 1% to 28% across the studies, and a pooled prevalence of severe disease of 25% [95% CI: 21-31%; I^2^=96%; n=36] (**Table 2**). The prevalence of death was 21% [95% CI: 18-24%; I^2^=78%; n=9] in the US and Europe, and 13% [95% CI: 9-18%; I^2^=93%; n=17] in China. Prevalence of severe disease was 18% [95% CI: 13-24%; I^2^=97%; n=10] for US and Europe, and 33% [95% CI: 27-41%; I^2^=95%; n=25] for China. The measures for severe disease are described in **Supplemental table 1**.

### Pooled Prevalence of Risk Factors and Association with Death or Severe Disease Age and Sex (Tables 2 and Figure 2)

Median age for people who died was 67 years [IQR: 63-71; I^2^=74%; n=18] and who had severe disease was 60 years [IQR: 58-63; I^2^=47%; n=26]. Seventy-six percent [95% CI: 71-83; I2=76%; n=7] of the deaths were in people aged ≥ 60 years and 68% [95% CI: 63-74; n=13] were in males. The CFR (95% CI) was 42% (34-50%) for age≥ 60 years and 30% (23-38%) for males. Patients aged≥ 60 years [summary relative risk (sRR): 3.77; 95% CI: 2.94-4.82; I^2^=73%; n=12] and males [sRR: 1.34; 95% CI: 1.23-1.54; I^2^=19%; n=17] had higher risk of death. The risk of severe disease was similarly higher (**Supplemental table 1**).

**Figure 2.**
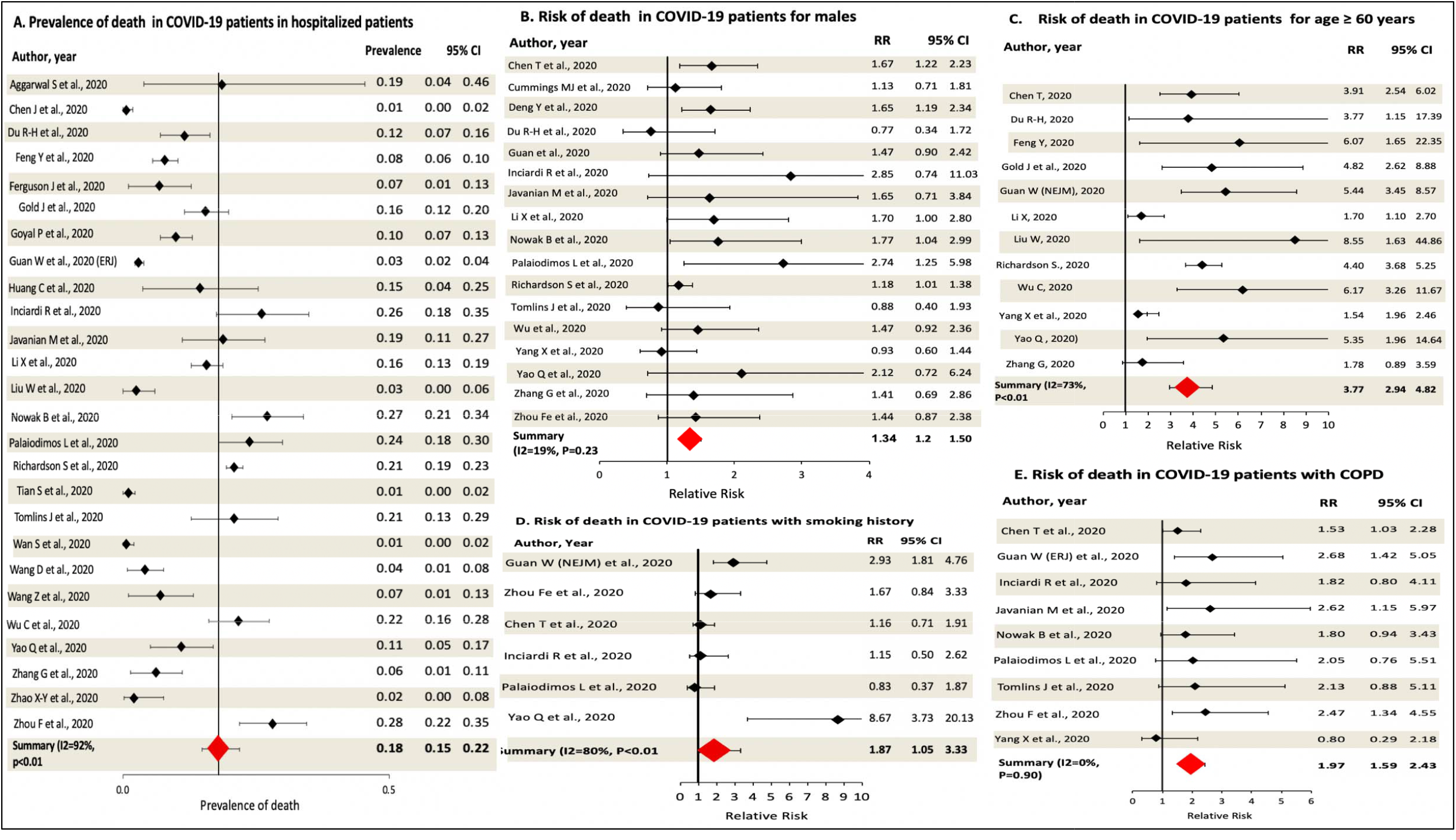
Prevalence of death in COVID-19 patients and association with epidemiological risk factors.

### Smoking and COPD (Table 2 and Figure 2)

The pooled prevalence for any history of smoking in the patients was 13% [95% CI: 11-16%; I^2^=22%; n=12]. For patients with smoking history, the CFR was 22% (95% CI: 11-42%) and CSR was 42% (95% CI: 33-53). Compared to never smokers, patients with smoking history had higher relative risk of death [sRR: 1.87; 95% CI: 1.05-3.33; I^2^=80%; n=6] and severe COVID-19 disease [sRR: 1.33; 95% CI: 1.16-1.54; I^2^=42%; n=15]. The prevalence of COPD was 8% [95% CI: 6-10%; I^2^=93%; n=27]. Patients with COPD had a CFR of 51% (95% CI: 36-71%), CSR of 49% (95% CI: 42-56%), a sRR of death of 2.80 [95% CI: 1.69-4.66; I^2^=82%; n=9].

### Hypertension (Tables 2 and Figure 3)

The pooled prevalence of hypertension in the COVID-19 patients was 47% [95% CI: 41-54% I^2^=98%; n=34], with a CFR in hypertensive patients of 24% (95% CI: 16-36%) and a CSR of 53% (95% CI: 44-64%). Of the COVID-19 patients that died, 63% [95% CI: 57-70%; I^2^=98%; n=13] were having hypertension. Hypertensives had higher relative risk of death [sRR: 1.84; 95% CI: 1.66-2.03; I^2^=34%; n=14] and severe disease [sRR: 1.62; 95% CI: 1.37-1.92; I^2^=79%; n=23] compared to non-hypertensives.

**Figure 3.**
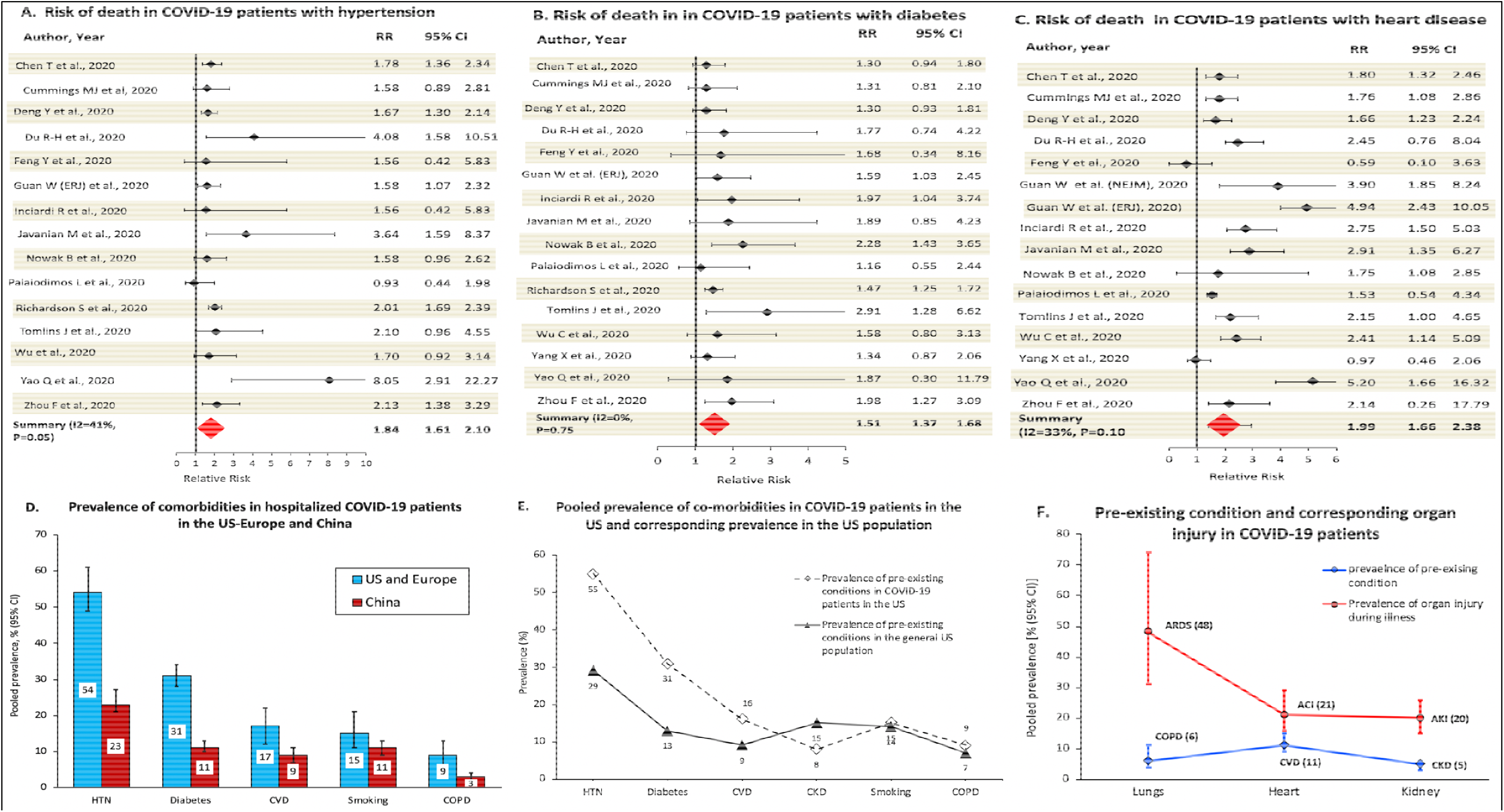
Risk of death in COVID-19 patients associated with pre-existing conditions, and prevalence of comorbidities and organ injury. HTN: hypertension; CVD: cardiovascular disease; COPD: chronic obstructive pulmonary disease; CKD: chronic kidney disease; AKI: acute kidney injury; ACI: acute cardiac injury; ARDS: acute respiratory distress syndrome

### Diabetes (Tables 2 and Figure 3)

The pooled prevalence of diabetes was 26% [95% CI: 22-30%; I^2^=96%; n=36] with a CFR of 22% (95% CI: 16-32%) and CSR of 46% (95% CI: 40-52%) in the diabetics. Of the COVID-19 patients that died, 38% [95% CI: 32-44%; I^2^=75%; n=13] were diabetics. Diabetics had higher relative risk of death [sRR: 1.51; 95% CI: 1.37-1.68; I^2^=0%; n=16] and severe disease [sRR: 1.50; 95% CI: 1.32-1.71; I^2^=65%; n=25] compared to non-diabetics.

### Cardiovascular Disease (Tables 2 and Figure 3)

The pooled prevalence of CVD was 14% [95% CI: 12-17%; I^2^=95%; n=35] with a CFR of 62% (95% CI: 48-78%) and CSR of 56% (95% CI: 47-68%) among cardiac patients. Of all the patients that died, 29% [95% CI: 21-41%; I^2^=81%; n=12] had heart disease. Cardiac patients had higher relative risk of death [sRR: 1.99; 95% CI: 1.72-2.38; I^2^=33%; n=16] and severe disease [sRR: 1.67; 95% CI: 1.42-1.96; I^2^=83%; n=20] compared to patients without cardiac disease.

### Chronic Kidney Disease (Tables 2 and Figure 3)

The pooled prevalence of CKD was 7% [95% CI: 5-10%; I^2^=93%; n=26] with a CFR of 49% (95% CI: 33-71%) and CSR of 34% (95% CI: 32-36%) in CKD patients. CKD was present in 23% [95% CI: 13-39%; I^2^=62%; n=7] of all COVID-19 patients that died. CKD patients had higher relative risk of death [sRR: 2.17; 95% CI: 1.30-3.13; I^2^=75%; n=8] and severe disease [sRR: 1.67; 95% CI: 1.30-2.16; I^2^=90%; n=14] compared to non-CKD patients.

### COVID Related Organ System Injury (Figure 3)

To understand how pre-existing health conditions may be correlated with the risk of specific organ injury, we calculated the pooled prevalence of acute injury to lung, heart and kidney for studies that reported prevalence of both the pre-existing condition(s) and corresponding organ injury. Pooled across 12 studies,^8,14,26,30,36-42,50^ the prevalence of COPD at baseline was 6% (95% CI: 4-11%) and that of ARDS was 48% (31-74%). The pooled prevalence of baseline CVD (n=11 studies) was 11% (95% CI: 9-12%) and that of acute cardiac injury (ACI) during hospitalization was 21% (95% CI: 16-29%).^4,14,26,30,31,35,37,38,40,50,53^ The prevalence of CKD (n=8 studies) was 5% (95% CI: 3-6%) and that of acute kidney injury (AKI) was 20% (95% CI: 15-26%).^4,14,26,30,36,37,43,50^

### Sensitivity and Sub-group Analyses

The relative risk of death in the patients was homogenously elevated for hypertension, diabetes, COPD and heart disease. We carried out sensitivity analyses by excluding outliers. For the risk of death for hypertension and smoking history, we removed the study by Yao et al^53^ which showed significantly higher risk compared to other studies; the results for both hypertension (sRR=2.02; 95% CI: 1.70-2.38) and smoking (sRR:1.59; 95% CI: 1.01-2.49) remained significant. Guan et al.^13^ had published a second study with additional patients and reported adjusted estimates for COPD, diabetes and hypertension. We used the adjusted risk estimates for the analyses. For the risk of death with other risk factors (CVD & CKD) for Guan et al., we conducted sensitivity analyses by using the counts only from the original study.^36^ The results [sRR (95% CI)] were similar as: CVD=1.99 (1.69-2.33) and CKD=1.90 (1.27-2.86). We observed variation in the prevalence of risk factors and death based on region. Upon sub-group analysis, we noted significantly higher prevalence of risk factors among COVID-19 patients in the US and Europe than in China (**Figure 3. D**). Pooled prevalence of co-morbidities between US-Europe and China differed as follows: **1) for US-Europe**: HTN=54% (95% CI: 49-61%); diabetes=31% (95% CI: 28-34); CVD=17% (12-22%); smoking history=15% (11-21%); COPD=9% (6-13%) and **2) For China**: HTN=23% (21-27%); diabetes=11% (10-13%); CVD=9% (7-11%); smoking history=11% (9-13%); and COPD=3% (2-4%).

### Publication Bias

We observed asymmetry in the funnel plot for studies that reported prevalence of death in COVID-19 patients (Egger’s test p=0.007 and Begg’s test p=004) (**Supplemental Figure 1**). On further analysis, the plot remained asymmetrical when restricted to studies from China (Egger’s p=0.060 and Begg’s p=0.053) but was symmetrical for studies from US-Europe (Egger’s p=0.462 and Begg’s p=0.210). We observed symmetrical funnel plots with no bias for pooled prevalence severe disease or for analyses of association of risk factors with death or severe disease.

## Discussion

We carried out a comprehensive systematic review and meta-analysis including 20594 hospitalized patients from 44 studies, 12591 from the US and Europe, and 7885 from China to investigate the prevalence and risk factors for death and severe disease in COVID-19 patients. We calculated a prevalence of death of 18%–21% for US and Europe, and 13% for China–and a prevalence of severe disease of 25%–18% for US and Europe, and 33% for China. In the COVID patients who died, risk factors were distributed as: age ≥ 60 years: 76%; males: 68%; hypertension: 63%; diabetes: 38%; heart disease: 29%; CKD: 23%; smoking history: 27%; and COPD: 12%.

In comparison with the overall prevalence of death of 18% for all COVID-19 hospitalized patients, the CFR was higher for patients with comorbidities: heart disease (62%), COPD (51%), CKD (49%), CLD (41%), hypertension (24%), diabetes (22%), and smoking history (22%). CFR was also higher for age≥ 60 years (42%) and for males (30%). The elevation in the risk of death was statistically significant for age ≥ 60 (sRR=3.8; 95% CI: 2.9-4.8), male sex 1.3 (95% CI: 1.2-1.5), smoking history (sRR=1.9; 95% CI: 1.1-3.3), COPD (sRR=2.0; 95% CI: 1.6-2.4), heart disease (sRR=2.0; 95% CI: 1.7-2.4), CKD (sRR=2.0; 95% CI: 1.3-3.1), hypertension (sRR=1.8; 95% CI: 1.7-2.0), and diabetes (sRR=1.5; 95% CI: 1.4-1.7). All of the risk factors we analyzed were positively associated with progression to severe disease as well (**Table 2 & 3**). The results suggest that older age, male sex and the co-morbidities increase the risk of progression to severe disease and death in COVID-19 patients.

While we are unsure of the reason behind significant tests for publication bias for prevalence of death in studies from China, the funnel plot showed several studies with lower prevalence of death. This may suggest under reporting of death for the several studies from China. While the lower median age as well as lower prevalence of co-morbidities for COVID-19 patients in China can explain the lower prevalence of death, this should not lead to a statistical bias in publication as observed. The lower prevalence of death is incommensurate with the higher prevalence of severe disease observed for the studies from China. China’s official death toll from COVID-19 was under-reported initially that was updated on April 17, 2020.^62^

There is ongoing debate regarding predisposition by cardiovascular disease– hypertension, diabetes, heart disease–toward increasing the risk of acquisition of SARS-CoV-2.^63-65^ On one hand, Angiotensin converting enzyme 2 (ACE2)–by blocking the renin angiotensin aldosterone system (RAAS) and decreasing or countering the vasoconstrictive, proinflammatory and profibrotic properties of Angiotensin-II through catalysis of ANG-II to Ang-(1-7)–exerts cardiovascular protective effects and can prevent acute lung injury from SARS-CoV-2.^63-65^ However, on the other hand, it is unclear if angiotensin converting enzyme inhibitors (ACEI) or angiotensin receptor blockers (ARB)–used for treatment of hypertension, heart disease and diabetes–increases susceptibility toward SARS-CoV-2 infection through potential upregulation of ACE2, the host protein used by SARS-CoV-2 as a co-receptor to translocate intracellularly.^63-66^ In this context, it would be reasonable to posit that a substantially higher prevalence of cardiovascular comorbidities in the hospitalized patients compared to the prevalence in the general population may suggest elevated risk of acquisition of SARS-CoV-2 for patients with cardiovascular risk factors. To this end, we found that the prevalence of hypertension (55%), diabetes (31%) and heart disease (16%) in the hospitalized COVID-19 patients in the US were substantially higher than the corresponding prevalence of hypertension (29%),^67^ diabetes (13%)^68^ and heart disease (9%)^15^ in the general US population (**Figure 3 E**), suggesting either a tendency for SARS-CoV-2 to more effectively establish infection in cardiovascular patients or to cause more severe disease among them prompting hospitalization. The prevalence of smoking history (15%), COPD (9%) and CKD (8%) in the COVID-19 patients in the US was similar to or lower than the population prevalence of smoking (14%),^69^ COPD (7%),^70^ and CKD (15%)^71^ in the country. In the prevalence of hypertension (23%), diabetes (11%), CVD (9%), smoking (11%) and COPD (3%) in the hospitalized patients in China (**Figure 3 D**) were all lower as compared to the US. The prevalence of these comorbidities in the COVID-19 patients was actually corresponding to the prevalence of hypertension (23%),^17^ diabetes (15%)^72^, and CVD (21%)^73^ in the general population of China. A previous meta-analysis also noted this observation.^18^ Notably, the prevalence of smoking (11%) and CVD (9%) in the hospitalized COVID-19 patients in China are inexplicably lower than the prevalence of smoking (15%) and CVD (16%) among hospitalized patients in the US, despite the prevalence of smoking (47% in Chinese males)^74^ and CVD (21%)^73^ in the general Chinese population being significantly higher than that of the US. As such, we are unsure whether the lower prevalence of comorbidities noted for the COVID-19 patients in China are representative of the true prevalence. There was a such great urgency and a race to publish data in the early phase of the outbreak. As such, there exists the possibility of substantial under-recording of data on covariables.

We assessed if patients with specific co-morbidities at baseline had higher risk of specific organ injury from SARS-CoV-2 during hospitalization. While the available data did not allow direct assessment of this relation, we compared the prevalence of comorbidities with the prevalence of corresponding organ system injury for studies that reported both baseline comorbidity and corresponding organ injury. We observed that the risk of acute lung injury (measured as ARDS) (48%), ACI (21%), and AKI (20%) were substantially higher the baseline prevalence of COPD (6%), heart disease (11%) and CKD (5%), respectively (**Figure 3 F**). The higher prevalence of acute organ injury than the prevalence of baseline comorbidity simply indicates that ARDS, ACI and AKI were also occurring in patients who did not have a corresponding comorbidity at baseline.

A limitation is that most studies simply reported frequencies of risk factor and did not present adjusted measures for disease severity or death. As such, the risk ratio we calculated from the frequencies are largely unadjusted estimates. Future studies could additionally present, at the least, age and sex adjusted measures for association of risk of comorbidities with death or severe disease. Many studies reported odds ratio for the measure of association between pre-existing conditions and risk of severe disease or death. Odds ratio poorly approximates risk ratio when the disease prevalence is high at baseline. For example, Zhou et al.^14^ calculated an odds ratio of 5.4 (95% CI: 0.96-30.4) for risk of death from COPD in COVID-19 patients whereas the risk ratio we calculated from the frequencies presented is RR=2.47; 95% CI: 1.34-4.55. Prevalence of severe disease or death in COVID-19 patients was high in several studies. Similarly, several meta-analyses calculated odds ratios instead of risk ratios to summarize the risk of disease severity or death in association with risk factors such as smoking, diabetes, hypertension and cardiovascular disease,^10,11,16^ often to be interpreted by media and even by researchers as a measure of relative risk. Lack of rigor in research design, analysis and interpretation could generate inconsistent and ungeneralizable results across studies leading to controversy and confusion around serious public health issues such as that existing for association (or not) of smoking with COVID-19 disease acquisition, severity or death.

As publications evolve at a pace that could be overwhelming for researchers and practitioners, we attempted to present a meaningful summary and inference for association of risk factors with death or severe disease from literatures published globally and an epidemiological framework for the risk of infection by SARS-CoV-2 based on presence of cardiovascular risk factors. This analysis can inform public health measures for COVID-19 screening and prevention, risk stratification and management of patients in clinical practice, analysis and presentation strategies for research data and inspire etiological investigations.

## Data Availability

All data are available.

## Research in Context

### Evidence before this study

COVID-19 is an ongoing pandemic. Prevalence of death as a result of COVID-19 has greatly varied across regions. Epidemiological risk factors for progression of COVID-19 to severe disease and death, and the risk of acquisition of SARS-CoV-2, the causal agent for COVID-19, based on presence of pre-existing conditions are insufficiently understood. Particularly, there exist controversy and gap in knowledge on how pre-existing cardiovascular disease including hypertension, diabetes, and heart disease and use of angiotensin converting enzyme inhibitors and angiotensin receptor blockers influence the risk of disease acquisition as well as disease progression.

### Added value of this study

Meta-analysis of 44 studies including 20285 COVID-19 patients revealed case fatality risk of 62% for heart disease, 51% for COPD, 49% for CKD, 24% for hypertension, 22% for diabetes, and 22% for smoking history. Of all patients that died, 76% were aged≥ 60 years, 68% were males, 63% had hypertension and 38% had diabetes. Age≥ 60, male sex, and pre-existing conditions were significantly association with death and severe disease. Also, the prevalence of cardiovascular risk factors–hypertension, diabetes, and heart disease–were substantially higher in COVID-19 patients than their prevalence in the general US population.

### Implications of all the available evidence

COVID-19 patients who are older, males and have pre-existing health conditions are at higher risk of progression to severe disease and death. The comparative prevalence data suggest people with cardiovascular risk factors may be at higher risk of acquisition of SARS-CoV-2 infection.

### Funding

Dr. Dorjee is supported by grants from private philanthropists; the Johns Hopkins Alliance for Healthier World (Grant # 80045453); National Institute of Allergy and Infectious Diseases of the National Institute of Health (Grant # K01AI148583); the United Nations STOP TB PARTNERSHIP TB REACH (Grant # 134126); and the Pittsfield Anti-tuberculosis Association (PATA).

## Notes

### Competing Interest Statement

The authors have declared no competing interest.

### Funding Statement

Dr. Dorjee is supported by grants from private philanthropists, Johns Hopkins Alliance for Healthier World (Grant # 80045453); the United Nations STOP TB PARTNERSHIP TB REACH (Grant # 134126); National Institute of Allergy and Infectious Diseases of the National Institute of Health (NIAID/NIH) (Grant # K01AI148583) and the Pittsfield Anti-tuberculosis Association (PATA). The content is solely the responsibility of the authors and does not represent the views of
the sponsors.

### Author Declarations

Meta-analysis study. IRB not relevant.

